# Increased extravascular lung water index (EVLWI) reflects rapid inflammatory oedema and mortality in COVID-19 associated ARDS

**DOI:** 10.1101/2020.09.11.20192526

**Authors:** Sebastian Rasch, Paul Schmidle, Sengül Sancak, Alexander Herner, Christina Huberle, Dominik Schulz, Ulrich Mayr, Jochen Schneider, Christoph Spinner, Fabian Geisler, Roland M. Schmid, Tobias Lahmer, Wolfgang Huber

## Abstract

**OBJECTIVE:** Nearly 5 % of the patients with COVID-19 develop an acute respiratory distress syndrome (ARDS). Extravascular lung water index (EVLWI) is a marker of pulmonary oedema which is associated with mortality in ARDS. In this study we evaluate whether EVLWI is higher in patients with COVID-19 associated ARDS as compared to controls and whether EVLWI has the potential to monitor disease progression.

**METHODS:** From the day of intubation, EVLWI, cardiac function were monitored by transpulmonary thermodilution in n=25 patients with COVID-19 and compared to a control group of 49 non-COVID-19 ARDS-patients.

**RESULTS:** EVLWI in COVID-19-patients was noticeably elevated and significantly higher than in the control group (17 (11-38) vs. 11 (6-26) mL/kg; p<0.001). High pulmonary vascular permeability index values (2.9 (1.0-5.2) versus 1.9 (1.0-5.2); p=0.003) suggest inflammatory oedema. By contrast, the cardiac parameters SVI, GEF and GEDVI were comparable. High EVLWI values were associated with viral persistence, prolonged intensive care treatment and mortality (23.2±6.7% vs. 30.3±6.0%, p=0.025).

**CONCLUSIONS:** Compared to the control group, COVID-19 results in markedly elevated EVLWI-values in patients with ARDS. EVLWI reflects a non-cardiogenic pulmonary oedema in COVID-19 associated ARDS and could serve as parameter to monitor ARDS progression.

## INTRODUCTION

COVID-19 is caused by Severe Acute Respiratory Coronavirus 2 (SARS-CoV-2) and shows a wide clinical spectrum covering asymptomatic cases, mild upper respiratory affectation, and severe pneumonia.^1,2^ While a majority of patients have a favorable outcome, higher age and underlying comorbidities are associated with a poor prognosis. Typically, patients with severe COVID-19 pneumonia suffer from dyspnea, hypoxemia, massive alveolar damage, progression to acute respiratory distress syndrome (ARDS) and multiple organ failure.^3^

The pathogenesis of the COVID-19 is poorly understood. As far as known, onset of COVID-19 associated ARDS leads to uncontrolled pulmonary inflammation, fluid accumulation, and progressive fibrosis that severely compromise oxygen and carbon dioxide exchange.^4^ Moreover, a complex immune response of the host versus the SARS-CoV-2 virus is assumed, which result in a liberation of soluble inflammatory proteins - a so called cytokine storm.^5-8^

In these most severely ill patients, computed tomography (CT) of the chest demonstrates an unprecedented, typical pattern which is suggestive to a degree that it is disease-defining, even if the SARS-CoV-2 PCR is negative.^9,10^

Regarding the predominantly higher age and a substantial prevalence of circulatory comorbidities such as coronary heart disease, peripheral artery disease, arterial hypertension and diabetes mellitus, the role of cardiogenic implications on pulmonary oedema has to be further studied.^2,3,5,7^

Single indicator transpulmonary thermodilution (TPTD) is a commercially available technology of advanced hemodynamic monitoring. TPTD provides bedside measurement of extravascular lung water index (EVLWI) which is a marker of pulmonary oedema. Additionally, crucial hemodynamics such as stroke volume index (SVI), global ejection fraction (GEF) and the preload marker global end-diastolic volume index (GEDVI) are derived from TPTD in parallel with EVLWI.^11^’^13^ Several studies demonstrated significant and independent association of EVLWI and its changes over time with mortality.^14^’^19^ A recent study in a non-COVID-19 cohort with ARDS patients suggests an improved and earlier prediction of 28-days-mortality compared to traditional scores of ARDS severity.^20^ Furthermore, TPTD-monitoring of critically ill non-COVID-19 patients was independently associated with a lower mortality in this study.

To date, in COVID-19-patients, there is a lack of data on hemodynamic key parameters, especially on EVLWI, generated by bedside TPTD.

Aim of our study is to investigate key hemodynamic and pulmonary parameters derived from TPTD in mechanically ventilated COVID-19-patients with ARDS compared to a recent non-COVID-19 cohort with ARDS.

## MATERIAL AND METHODS

The study protocol was approved by the Institutional Review Board (Ethics committee of Technical University of Munich; Approval No. 178/20S) as essential part of the register study CORRECT: COVID Registry REChts der Isar intensive care Trial. The study was registered at the Clinical Trial Registry (No. (ISRCTN10077335). Additional data of the study and control group is reported in supplementary digital table 1.

**Table 1:**
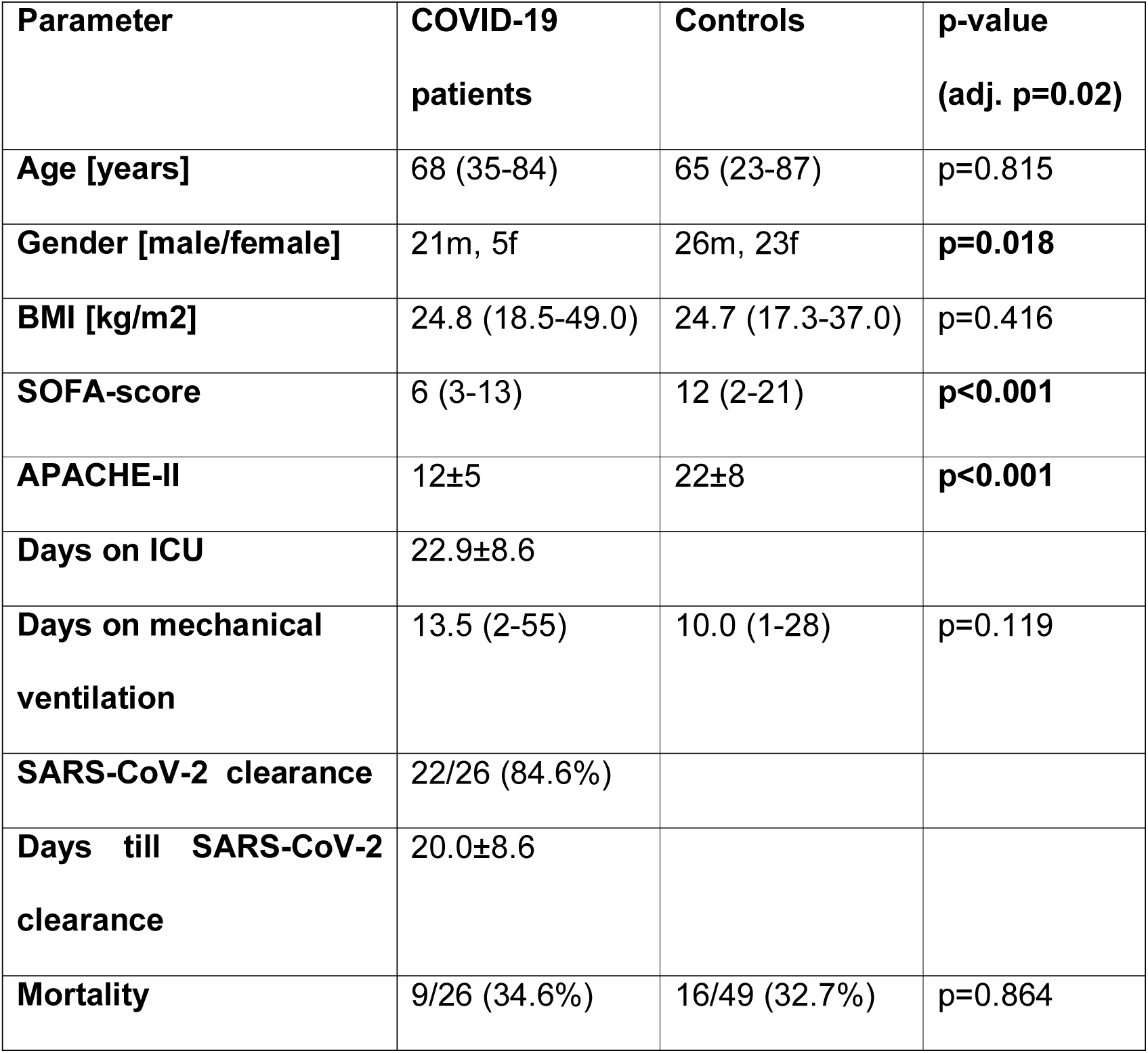
Patient characteristics. (BMI: body mass index, ICU: intensive care unit)

All patients or their legal representatives gave written informed consent. The study was conducted in a COVID-19-ICU of a university hospital with 14 beds.

### Inclusion and exclusion criteria

In addition to the diagnosis of COVID-19, confirmed by a positive SARS-CoV-2-PCR, all patients were intubated, mechanically ventilated, and suffered from a documented ARDS, according to the Berlin definition.^21^ Patients were excluded, if TPTD was contra-indicated (lower extremity peripheral artery disease grade II or above according to the Forestier classification) or not feasible within the first 12 hours after intubation. Since extracorporeal membrane oxygenation might lead to incorrect measurement of EVLWI and GEDVI, TPTD measurements during extracorporeal membrane oxygenation (ECMO) were not included.^22^

According to the local standard TPTD was performed at least once within 24h as described previously.^13,23^

In brief a, 5F thermistor-tipped arterial line (PV2025L20, Pulsiocath, Pulsion Medical Systems, SE Feldkirchen Germany) was inserted into the femoral artery. The thermistor line and the pressure line of the arterial catheter as well as a second thermistor on the central venous catheter for measurement of the injectate temperature were connected to the hemodynamic monitor (PiCCO-2 (8 beds) or PulsioFlex (6 beds), both Pulsion Medical Systems, SE Feldkirchen Germany). The TPTD-curve was registered and analyzed after injection of 15 mL icecold 0.9% saline solution via the central venous catheter (CVC). Each TPTD measurement represents the mean of three consecutive thermodilution measurements within 5 minutes.

EVLWI was indexed to predicted bodyweight as suggested by the manufacturer.^24^ To derive EVLWI, GEDVI, SVI, GEF and all other parameters provided by the PiCCO, we used the most recent software V3.1, which corrects GEDVI for *femoral* CVC indicator injection.^25^ Since this correction does not pertain to pulmonary vascular permeability index (PVPI), in both cohorts PVPI_fem derived from femoral indicator injection was corrected as suggested recently.^26^ Correction is based on two formulas:

PVPI_fem_corrected = PVPI_fem * GEDVI_fem_uncorrected/GEDVI_fem_corrected

and

GEDVI_fem_corrected = 0.539 *GEDVI_fem_uncorrected -15.15 + 24.49 *CI_fem + 2.311*IBW ^25,26^

PVPI_fem, GEDVI_fem and CI_fem: PVPI, GEDVI and PVPI derived from femoral indicator injection. IBW: Ideal bodyweight. Calculation see^16^.

### Primary endpoint

Comparison of EVLWI in COVID-19-ARDS-patients with a recent non-COVID-19- cohort with ARDS.^20^ The non-COVID-19-cohort comprised 49 consecutive patients with TPTD-monitoring and ARDS.^21^ All patients of this cohort were treated in the same ICU as the COVID-19-patients and had been treated before 2019 (see Clinical Study Registration No. ISRCTN32938630; Institutional Review Board (Ethics committee of Technical University of Munich), Approval No. 343/18 S).

### Secondary endpoints

- EVLWI as potential parameter to monitor ARDS progression

- SVI, GEF and GEDVI in ARDS patients with and without COVID-19

### Power calculation

Based on two independent study groups, a continuous endpoint (EVLWI with a mean of 12.5±4.9mL/kg in the control cohort p-value and an estimated EVLWI of 18±7mL/kg in the COVID-19-cohort, a number of 49 controls and 25 COVID-19-patients would result in a statistical power of >90% with a p-value of p<0.05.^20^

### Statistics

Statistical analysis was performed using IBM SPSS Statistics 25 (SPSS Inc, Chicago, Illinois, USA). Samples were checked for normal distribution using the Shapiro-Wilk test. Descriptive data of normally distributed parameters were presented as mean ± standard deviation and as median and range for non-parametric parameters. The Mann-Whitney-U and Kruskal-Wallis tests were used to analyze non-parametric variables and the t-test and a one-way analysis of variances (ANOVA) to analyze variables with normal distribution. To compare qualitative parameters, chi-square test and in small samples (expected frequency of test variable less than 5) Fisher’s exact test was used. All statistical tests were two-sided with a level of significance (p-value) of 5%. Multivariate linear regression models were used to identify parameters that are independently associated with higher EVLWI and PVPI values. Factors with a significant p-value below 0.05 in univariate analysis were included in the regression models. To control the false discovery rate after multiple testing we adjusted the level of significance by the Benjamini-Hochberg procedure.

## RESULTS

In total 74 patients with ARDS were included in the study (25 with COVID-19 and 49 without). Patient characteristics are shown in Table 1.

### Biometric data and scores

Patients with COVID-19 were more frequently male compared to the controls (20/25 (80%) vs. 26/49 (53%); p=0.041; Table 1). SOFA and APACHE-II-score were higher in the COVID negative control group.

#### Respiratory data

The respiratory data is reported in table 2.

Summarizing several of the respiratory parameters, the oxygenation index (OI = Paw_mean * FiO_2_/pO_2_) was 66% higher in the COVID-19-cohort (14.1 ±9.9 vs. 8.5±4.4; p=0.005).

**Table 2:** Respiratory baseline parameters. (* mild vs. moderate/severe, P_peak: maximal inspiratory pressure, PEEP: positive end-expiratory pressure, pO2: partial pressure of oxygen, OI: oxygenation index, ARDS: acute respiratory distress syndrome)

| Parameter | COVID-19 patients | Controls | p-value |
| --- | --- | --- | --- |
| **P_peak** | 26 (20-40) | 27.5 (12-32) | $p=0.631$ |
| **PEEP [cm H<sub>2</sub>O]** | 14 (5-20) | 8 (6-15) | **$p<0.001$** |
| **Tidal volume [mL]** | $530 \pm 110$ | $494 \pm 148$ | $p=0.245$ |
| **pO<sub>2</sub>/FiO<sub>2</sub> (Horovitz-index)** | $148 \pm 81$ | $187 \pm 62$ | **$p=0.041$** |
| **OI** | $14.1 \pm 9.9$ | $8.5 \pm 4.4$ | **$p=0.005$** |
| **ARDS Berlin-definition** |  |  |  |
| - Mild | 5/26 (19%) | 22/49 (45%) | **$p=0.028^*$** |
| - Moderate | 16/26 (62%) | 24/49 (49%) |  |
| - severe | 5/26 (19%) | 3/49 (6%) |  |

### Parameters derived from TPTD and pulse contour analysis (PCA)

TPTD data is reported in table 3.

**Table 3:**
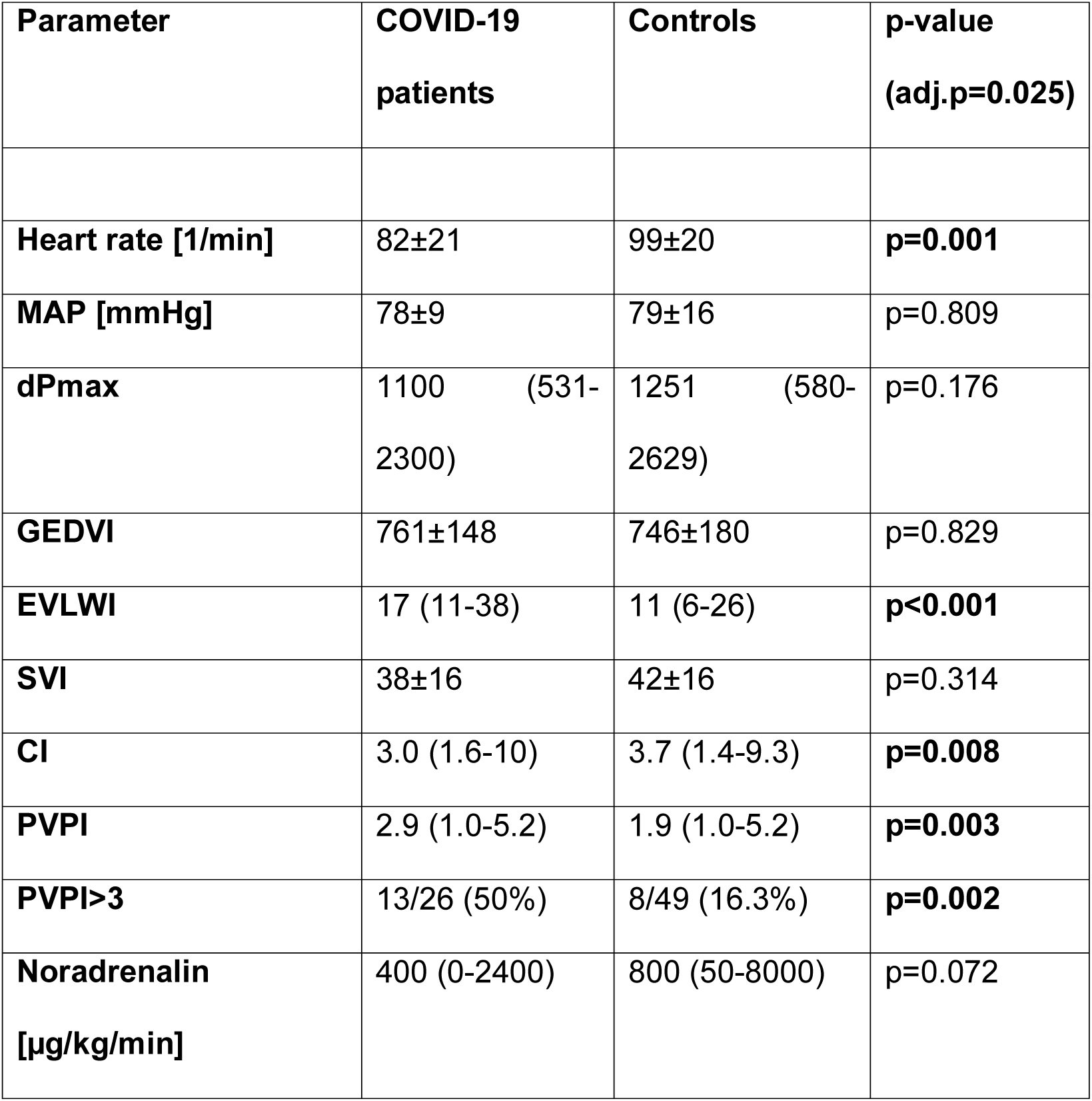
Hemodynamic data: 1^st^ measurement. (MAP: mean arterial pressure, dPmax: cardiac contractility index, GEDVI: global end-diastolic volume index, EVLWI: extra vascular lung water index, SVI: stroke volume index, CI: cardiac index, PVPI: pulmonary vascular permeability index)

| Parameter | COVID-19 patients | Controls | p-value (adj.p=0.025) |
| --- | --- | --- | --- |
| Heart rate [1/min] | 82±21 | 99±20 | **p=0.001** |
| MAP [mmHg] | 78±9 | 79±16 | p=0.809 |
| dPmax | 1100 (531-2300) | 1251 (580-2629) | p=0.176 |
| GEDVI | 761±148 | 746±180 | p=0.829 |
| EVLWI | 17 (11-38) | 11 (6-26) | **p<0.001** |
| SVI | 38±16 | 42±16 | p=0.314 |
| CI | 3.0 (1.6-10) | 3.7 (1.4-9.3) | **p=0.008** |
| PVPI | 2.9 (1.0-5.2) | 1.9 (1.0-5.2) | **p=0.003** |
| PVPI>3 | 13/26 (50%) | 8/49 (16.3%) | **p=0.002** |
| Noradrenalin [µg/kg/min] | 400 (0-2400) | 800 (50-8000) | p=0.072 |

### EVLWI and pulmonary vascular permeability index (PVPI) in COVID-19 patients versus controls

EVLWI on day-1 (the day of intubation) and the highest EVLWI within the first 14 days after intubation (25.0 (15.0-43.0) vs. 14.0 (7.0-54.0); p<0.001) were substantially higher in COVID-19-patients vs. pp-COVID-19-patients (Table 3; Fig. 1 BOXPLOTS).

**Figure 1:**
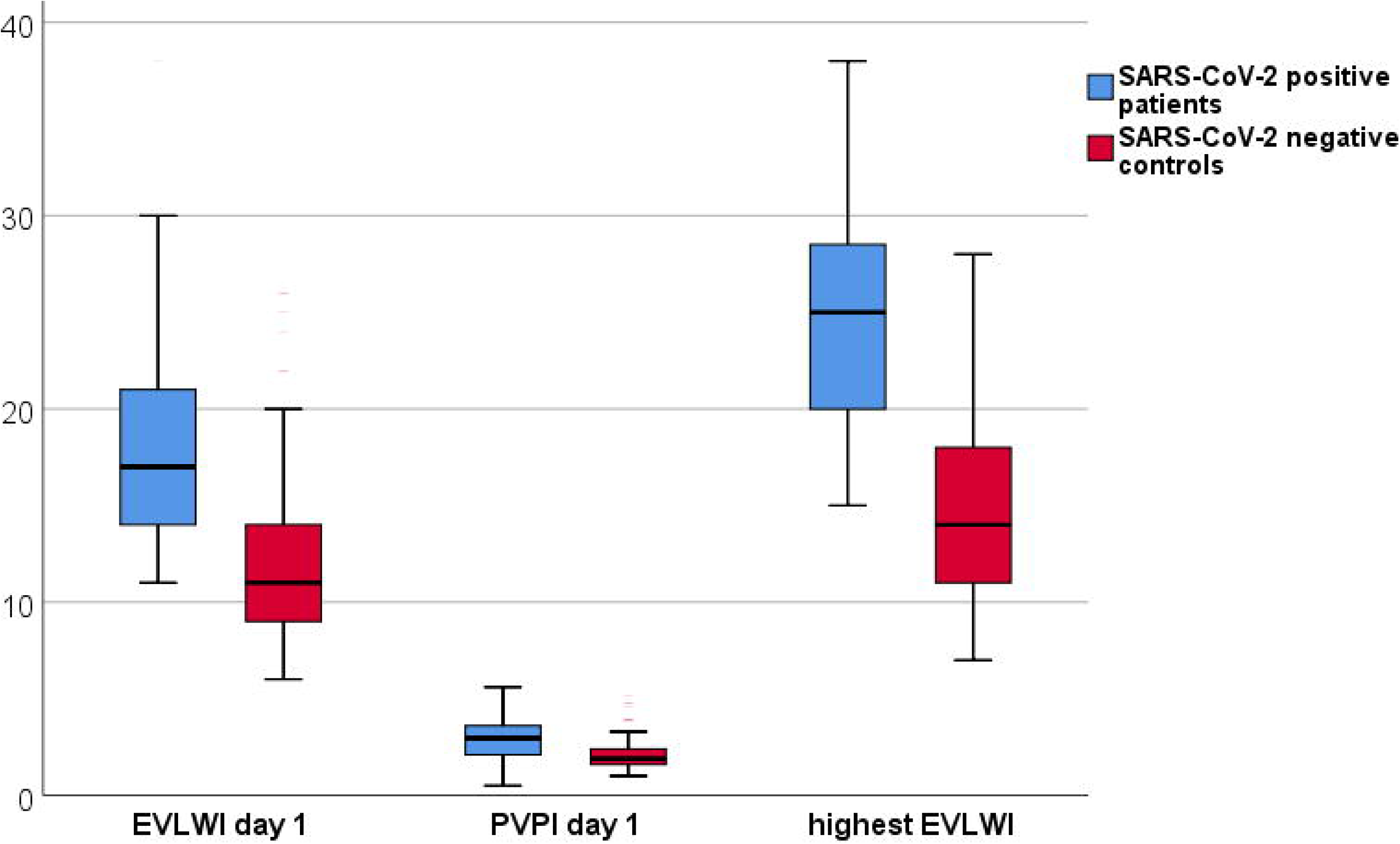
Boxplots comparing extra vascular lung water index (EVLWI) and pulmonary vascular permeability index (PVPI) on day 1 and highest EVLWI between patients with and without COVID-19

High PVPI values (≥3) are associated with inflammation and pulmonary origin, whereas low values are in line with cardiogenic or mixed pulmonary oedema. PVPI is calculated as a ratio from unindexed extravascular lung water EVLW divided by pulmonary blood volume PBV. PBV is assumed to be about 25% of unindexed GEDV (PVPI = EVLW/(0.25*GEDV)).^26^

PVPI was significantly higher in patients with COVID-19 compared to the controls on day-1 (Table 2 and Fig. 1).

In univariate analysis, the first EVLWI was associated with COVID-19 (r=0.503; p<0.001) and correlated with low body mass index (BMI) (r=-0.264; p=0.035), but not to gender, height, age, heart rate, MAP, SVRI, CVP, GEDVI, dPmax, SVI, CI, CPI and noradrenalin dosage.

Multivariate regression analysis (r=0.508; R^2^=0.258) regarding EVLWI including COVI-19 yes/no and BMI demonstrated that both COVID-19 (p<0.001, T=-4.801) and BMI (p=0.03, T=-2.214) were independently associated with higher EVLWI values.

PVPI was univariately associated with COVID-19, low BMI (r=-0.338; p=0.003) and low SVI (r=-0.230; p=0.048), but not with gender, height, age, heart rate, MAP, SVRI, CVP, dPmax, SVI, GEF, CPI and noradrenalin dosage.

In multivariate analysis (r=0.519; R2=0.269), PVPI was independently associated with COVID-19 (p=0.001; T=-3.481) and low BMI (p=0.001, T=-3.422), but not with SVI. GEDVI and EVLWI were not included in the multivariate analysis regarding PVPI, since PVPI is derived from the ratio of unindexed EVLW divided by 0.25*GEDV.A significant EVLWI decrease during the first days of mechanical ventilation was inversely associated with discharge from intensive care unit after less than 14 days and mortality (EVLWI at day 10 after intubation: 19.2±7.5 vs. 10.0±1.4, p=0.002, figure 2; mortality: deltaEVLWI 7 (0-22) versus 3 (0-12), p=0.021). Persistence of positive SARS-CoV-2 samples in COVID-19 patients during the ICU stay was associated with mortality (17/17 vs. 5/9, p=0.008). The highest EVLWI is associated with SARS-CoV-2 clearance (29.7±2.5 vs. 24.6±7.4, p=0.046) and mortality (23.2±6.7 vs. 30.3±6.0, p=0.025). The highest EVLWI was measured 5.2±4.4 days after intubation in the patients with COVID-19 associated ARDS.

**Figure 2:**
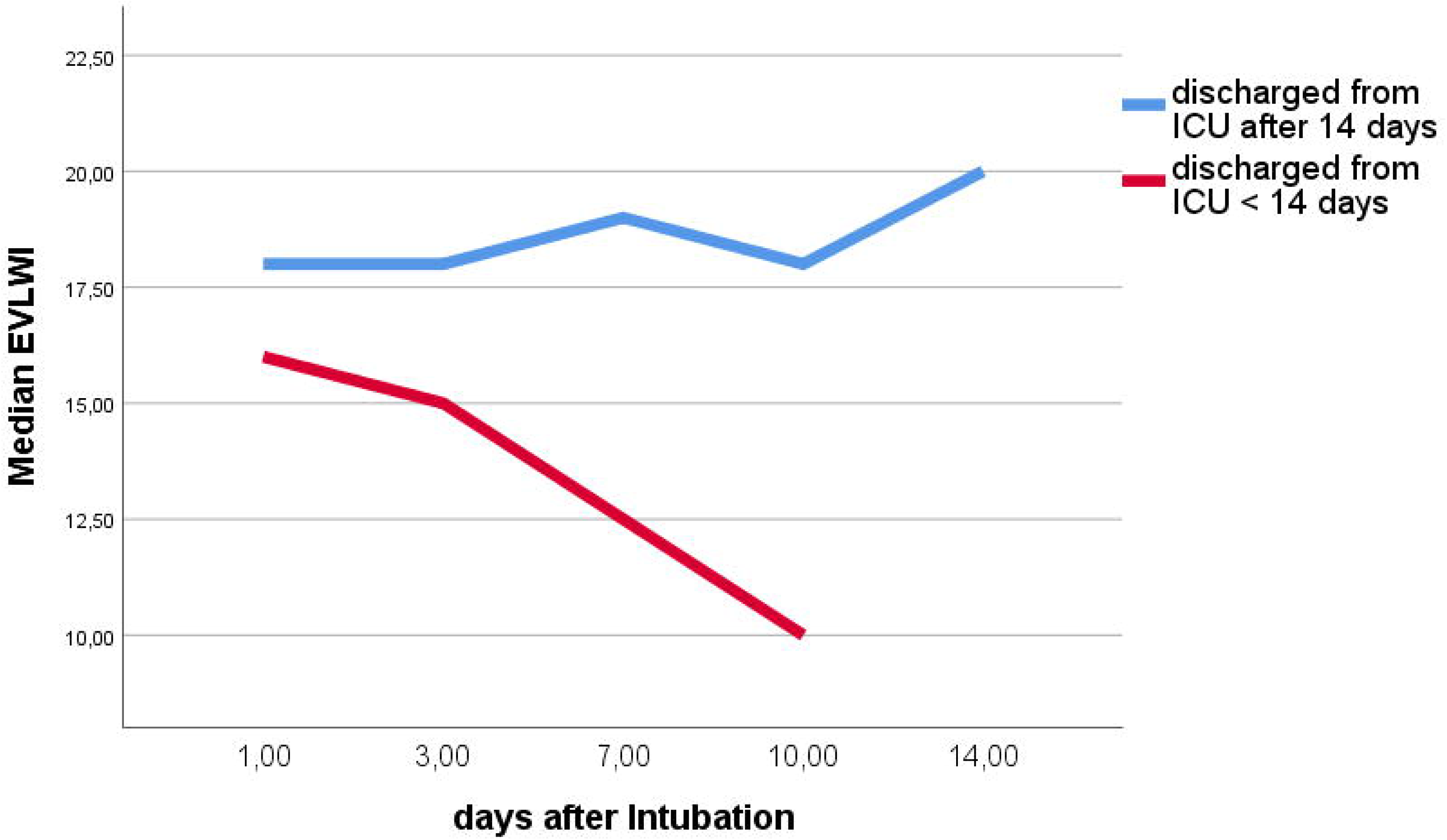
Extra vascular lung water index (EVLWI) of patients with COVID-19 who required less and more than 14 days of treatment on intensive care unit (ICU)

### Preload markers GEDVI and CVP

By contrast, the static preload markers GEDVI (761 ±168 vs. 749±180 mL/m^2^; p=0.882; Fig. 3) and CVP (16.4±7.4 vs. 17.9±8.0 mmHg; p=0.446) were not significantly different between ARDS-patients with and without COVID-19.

### Parameters of cardiac function in COVID-19 patients versus controls

Global ejection fraction (GEF) (20.9±6.0 vs. 23.8±7.3 %; p=0.098; Fig. 3) and stroke volume index (SVI) and dPmax were comparable for patients with and without COVID-19 on day-1 (table 2 and supplementary table 1).

## DISCUSSION

Symptoms of SARS-CoV-2 infections range from asymptomatic to severe ARDS. Similarly, there are patients with COVID-19 associated ARDS that recover within several days and others that require mechanical ventilation for weeks or do not recover at all. The reasons for this discrepancy are unclear and it is often hard to estimate an individual patients’ prognosis. According to published data a high EVLWI is associated with mortality in patients with ARDS.^15-17^ A median EVLWI of 17 ml/kg is not only higher than in our previous non-COVID-19 ARDS cohort, but also compared to previous studies in patients with ARDS.^14,16-19^ The absolute, nonindexed EVLW for a 70kg healthy patient would be around 500mL. In non COVID-19 ARDS patients it is 900 mL with the best cut-off to predict increased mortality at 1000mL.^27^ In COVID-19 patients EVLW reaches up to 2600 mL. Hence, there is no defined EVLWI cut off for the prediction of mortality as absolute EVLWI values are not comparable between patients with SARS-CoV-2 induced ARDS and with ARDS by other causes. While mortality is the same in both groups, EVLWI values differ significantly. In conclusion, high EVLWI values compared to other patients with COVID-19 predict mortality. In addition, the course of EVLWI values can help to monitor respiratory function of COVID-19 patients. Decreasing EVLWI values were associated with improved respiration and consequently shorter time on ICU. The high EVLWI values in patients with COVID-19 reflect the extent of pulmonary oedema caused by this viral pneumonia.

This degree of rapid inflammation is so far only known in severe acute pancreatitis.^28^ The morphologic correlate of pronounced pulmonary inflammation appears as diffuse interstitial oedema on CT that can affect large parts of the pulmonary tissue.^10^ A recent autopsy study reported pronounced endothelial damage and widespread capillary microthrombi in COVID-19 ARDS.^29^ Similar to Sepsis, a massive inflammatory response might explain this microangiopathy. In combination with intravascular coagulation and capillary leakage this results in extensive pulmonary oedema. Lungs of COVID-19-patients with ARDS have a lower weight at autopsy compared to influenza associated ARDS, which seems contrary to the increased EVLWI values. However, these two findings might be due to the different time point when measurements were performed. EVLWI values derived from the first days after intubation whereas autopsy is carried out later after termination of treatment. Interestingly, in multivariate regression analysis EVLWI was negatively associated with BMI, whereas recent reports e.g. from Italy suggest body weight to be associated with a bad outcome in COVID-19.^30^ This might be explained by the fact that EVLWI is indexed to predicted bodyweight. There is data supporting an indexation to height rather than body weight as height increases EVLWI values and an EVLW indexed to height predicts FiO2/pO2 more accurately than an EVLW indexed to ideal body weight.^16,24^ Increasing height on the other hand results in lower BMI values which might cause the negative association of BMI and EVLWI.

In addition to the absolute increase in EVLWI, our study gives several hints that this pulmonary oedema is mainly non-cardiogenic. The PiCCO-device combines TPTD with pulse contour analysis and provides a number of well-validated parameters of cardiac function. To facilitate decision support, a number of ratios is calculated, including PVPI and GEF (GEF = 4*stroke volume divided by GEDV).

PVPI relates EVLWI to preload (PVP = EVLW/(0.25*GEDV)). High values (in particular above 3) indicate pulmonary origin of the oedema with a normal GEDV. By contrast, elevated EVLWI-values in the context of a PVPI <2 suggests cardiac dilatation with an elevated GEDVI. A PVPI of 3.1 ±1.3 in our COVID-19-cohort is clearly in line with an inflammatory origin of the pulmonary oedema.

This is further supported by GEF of 21 ±6%, SVI of 38±17mL/m2 and dPmax of 1133±402 mmHg/s. These parameters were comparable between COVID-19- and non-COVID-19-patients in our study. Mean values of GEF, SVI and dPmax were slightly below the normal range. However, these normal ranges are given for a population with a representative age distribution. A recent study demonstrated that cardiac function as measured by cardiac output (CO) substantially decreases with older age (independent decrease of CO of 66mL/min per year).^31^ Therefore, GEF, SVI and dPmax might be considered within the age-adjusted normal range.

Repeated CT scans are an alternative diagnostic tool to monitor inflammation and ARDS progression. But inter-observer agreement depends on experienced staff and transport of ventilated patient always inherits a risk for the patient.^32^ As demonstrated in our study TPTD is a bedside available method to directly measure EVLWI with a limited invasiveness in the ICU-setting. It has been well validated compared to the more invasive double-indicator technique.^27,33,34^ EVLWI has not only the potential to predict mortality but also to monitor ARDS and the extent of pulmonary oedema during intensive care treatment.

### Limitations of the study

Due to its design as a mono-center study a selection bias might be discussed.

Slight baseline differences of the biometric data from COVID-19 and control cohorts can most likely be explained by older age and predominantly male gender in the COVID-19-cohort.

## CONCLUSION

EVLWI values in COVID-19 patients with ARDS are significantly higher than in the comparable control group. High EVLWI values are associated with increased mortality in patients with COVID-19 associated ARDS. EVLWI reflects a non-cardiogenic pulmonary oedema in COVID-19 associated ARDS and might serve as parameter to monitor ARDS progression.

## Data Availability

All data relevant for the analysis and conclusions of this study are included in this published article (and its Supplementary Information files). Exceeding information is available from the corresponding author on reasonable request.

## Author contributions

SR, PS, JS, CS, FG, RMS, TL and WH contributed to the design of the study.

SR, SS, AH, CH, DS, UM and TL were responsible for data collection.

SR, PS, TL and WH analysed the data.

SR and WH drafted the manuscript, all authors edited this and approved its final version.

## Supplementary data

Supplementary table 1: Additional respiratory and hemodynamic parameters

## REFERENCES

1 Huang, C. et al. Clinical features of patients infected with 2019 novel coronavirus in Wuhan, China. Lancet 395, 497–506, doi:10.1016/S0140-6736(20)30183-5 (2020).

2 Zhou, F. et al. Clinical course and risk factors for mortality of adult inpatients with COVID-19 in Wuhan, China: a retrospective cohort study. Lancet 395, 1054–1062, doi:10.1016/S0140-6736(20)30566-3 (2020).

3 Wu, C. et al. Risk Factors Associated With Acute Respiratory Distress Syndrome and Death in Patients With Coronavirus Disease 2019 Pneumonia in Wuhan, China. JAMA Intern Med, doi:10.1001/jamainternmed.2020.0994 (2020).

4 Yi, Y., Lagniton, P. N. P., Ye, S., Li, E. & Xu, R. H. COVID-19: what has been learned and to be learned about the novel coronavirus disease. Int J Biol Sci 16, 1753–1766, doi:10.7150/ijbs.45134 (2020).

5 Mo, P. et al. Clinical characteristics of refractory COVID-19 pneumonia in Wuhan, China. Clin Infect Dis, doi:10.1093/cid/ciaa270 (2020).

6 Spina, S. et al. The response of Milan’s Emergency Medical System to the COVID-19 outbreak in Italy. Lancet 395, e49-e50, doi:10.1016/S0140-6736(20)30493-1 (2020).

7 Wang, D. et al. Clinical Characteristics of 138 Hospitalized Patients With 2019 Novel Coronavirus-Infected Pneumonia in Wuhan, China. JAMA, doi:10.1001/jama.2020.1585 (2020).

8 Li, G. et al. Coronavirus infections and immune responses. J Med Virol 92, 424–432, doi:10.1002/jmv.25685 (2020).

9 Ai, T. et al. Correlation of Chest CT and RT-PCR Testing in Coronavirus Disease 2019 (COVID19) in China: A Report of 1014 Cases. Radiology, 200642, doi:10.1148/radiol.2020200642 (2020).

10 Li, K. et al. CT image visual quantitative evaluation and clinical classification of coronavirus disease (COVID-19). Eur Radiol, doi:10.1007/s00330-020-06817-6 (2020).

11 Combes, A., Berneau, J. B., Luyt, C. E. & Trouillet, J. L. Estimation of left ventricular systolic function by single transpulmonary thermodilution. Intensive Care Med 30, 1377–1383, doi:10.1007/s00134-004-2289-2 (2004).

12 Perny, J., Kimmoun, A., Perez, P. & Levy, B. Evaluation of cardiac function index as measured by transpulmonary thermodilution as an indicator of left ventricular ejection fraction in cardiogenic shock. Biomed Res Int 2014, 598029, doi:10.1155/2014/598029 (2014).

13 Huber, W. et al. Volume assessment in patients with necrotizing pancreatitis: a comparison of intrathoracic blood volume index, central venous pressure, and hematocrit, and their correlation to cardiac index and extravascular lung water index. Crit Care Med 36, 2348–2354, doi:10.1097/CCM.0b013e3181809928 (2008).

14 Brown, L. M. et al. Comparison of thermodilution measured extravascular lung water with chest radiographic assessment of pulmonary oedema in patients with acute lung injury. Ann Intensive Care 3, 25, doi:10.1186/2110-5820-3-25 (2013).

15 Craig, T. R. et al. Extravascular lung water indexed to predicted body weight is a novel predictor of intensive care unit mortality in patients with acute lung injury. Crit Care Med 38, 114–120, doi:10.1097/CCM.0b013e3181b43050 (2010).

16 Huber, W. et al. Association between different indexations of extravascular lung water (EVLW) and PaO2/FiO2: a two-center study in 231 patients. PLoS One 9, e103854, doi:10.1371/journal.pone.0103854 (2014).

17 Jozwiak, M. et al. Extravascular lung water is an independent prognostic factor in patients with acute respiratory distress syndrome. Crit Care Med 41, 472–480, doi:10.1097/CCM.0b013e31826ab377 (2013).

18 Tagami, T. et al. Early-phase changes of extravascular lung water index as a prognostic indicator in acute respiratory distress syndrome patients. Ann Intensive Care 4, 27, doi:10.1186/s13613-014-0027-7 (2014).

19 Zhao, Z. et al. Prognostic value of extravascular lung water assessed with lung ultrasound score by chest sonography in patients with acute respiratory distress syndrome. BMC Pulm Med 15, 98, doi:10.1186/s12890-015-0091-2 (2015).

20 Huber, W. et al. Prediction of Outcome in Patients With ARDS: A Prospective Cohort Study Comparing ARDS-definitions and Other ARDS-associated Parameters, Ratios and Scores at Intubation and Over Time. PloS one 15, doi:10.1371/journal.pone.0232720 (2020).

21 Ranieri, V. M. et al. Acute respiratory distress syndrome: the Berlin Definition. JAMA 307, 2526–2533, doi:10.1001/jama.2012.5669 (2012).

22 Herner, A. et al. Transpulmonary thermodilution before and during veno-venous extra-corporeal membrane oxygenation ECMO: an observational study on a potential loss of indicator into the extra-corporeal circuit. J Clin Monit Comput, doi:10.1007/s10877-01900398-6 (2019).

23 Hofkens, P. J. et al. Common pitfalls and tips and tricks to get the most out of your transpulmonary thermodilution device: results of a survey and state-of-the-art review. Anaesthesiol Intensive Ther 47, 89–116, doi:10.5603/AIT.a2014.0068 (2015).

24 Huber, W. et al. Extravascular lung water and its association with weight, height, age, and gender: a study in intensive care unit patients. Intensive Care Med 39, 146–150, doi:10.1007/s00134-012-2745-3 (2013).

25 Saugel, B. et al. Transpulmonary thermodilution using femoral indicator injection: a prospective trial in patients with a femoral and a jugular central venous catheter. Crit Care 14, R95, doi:10.1186/cc9030 (2010).

26 Huber, W. et al. Comparison of pulmonary vascular permeability index PVPI and global ejection fraction GEF derived from jugular and femoral indicator injection using the PiCCO-2 device: A prospective observational study. PLoS One 12, e0178372, doi:10.1371/journal.pone.0178372 (2017).

27 Sakka, S. G. et al. Assessment of cardiac preload and extravascular lung water by single transpulmonary thermodilution. Intensive Care Med 26, 180–187, doi:10.1007/s001340050043 (2000).

28 Iyer, H., Elhence, A., Mittal, S., Madan, K. & Garg, P. K. Pulmonary complications of acute pancreatitis. Expert Rev Respir Med 14, 209–217, doi:10.1080/17476348.2020.1698951 (2020).

29 Ackermann, M. et al. Pulmonary Vascular Endothelialitis, Thrombosis, and Angiogenesis in Covid-19. N Engl J Med, doi:10.1056/NEJMoa2015432 (2020).

30 Giacomelli, A. et al. 30-day mortality in patients hospitalized with COVID-19 during the first wave of the Italian epidemic: a prospective cohort study. Pharmacol Res, 104931, doi:10.1016/j.phrs.2020.104931 (2020).

31 Saugel, B. et al. Indexation of cardiac output to biometric parameters in critically ill patients: A systematic analysis of a transpulmonary thermodilution-derived database. J Crit Care 30, 957–962, doi:10.1016/j.jcrc.2015.06.011 (2015).

32 Sauter, A. W. et al. Intraobserver and interobserver agreement of volume perfusion CT (VPCT) measurements in patients with lung lesions. Eur J Radiol 81, 2853–2859, doi:10.1016/j.ejrad.2011.06.047 (2012).

33 Katzenelson, R. et al. Accuracy of transpulmonary thermodilution versus gravimetric measurement of extravascular lung water. Crit Care Med 32, 1550–1554, doi:10.1097/01.ccm.0000130995.18334.8b (2004).

34 Tagami, T. et al. Validation of extravascular lung water measurement by single transpulmonary thermodilution: human autopsy study. Crit Care 14, R162, doi:10.1186/cc9250 (2010).

